# Monkeypox caused less worry than COVID-19 among the general population during the first month of the WHO Monkeypox alert

**DOI:** 10.1101/2022.07.07.22277365

**Authors:** Mohamad-Hani Temsah, Fadi Aljamaan, Shuliweeh Alenezi, Khalid Alhasan, Basema Saddik, Ahmad Al-Barag, Ali Alhaboob, Nezar Bahabri, Fatimah Alshahrani, Abdulkarim Alrabiaah, Ali Alaraj, Feras Bahkali, Khaled Alkriadees, Amr Jamal, Rabih Halwani, Fahad AlZamil, Sarah Al-Subaie, Mazin Barry, Ziad A Memish, Jaffar A. Al-Tawfiq

## Abstract

**Background:** Monkeypox re-emerged in May 2022 as another global health threat. This study assessed the public’s perception, worries, and vaccine acceptance for Monkeypox and COVID-19 during the first month of WHO announcement.

**Methods:** A national cross-sectional survey was conducted between May 27 and June 5, 2022, in Saudi Arabia. Data were collected on sociodemographic characteristics, previous infection with COVID-19, worry levels regarding Monkeypox compared to COVID-19, awareness, and perceptions of Monkeypox, and vaccine acceptance.

**Results:** Among the 1546 participants, most respondents (62%) were more worried about COVID-19 than Monkeypox. Respondents aged 45 years and above and those with a university degree or higher had lower odds of agreement with Monkeypox vaccination (OR .871, p-value .006, OR .719, p-value <0.001), respectively. Respondents with moderate to a high level of self and family commitment to infection control precautionary measures and those who expressed self and family worry of Monkeypox infection had significantly higher odds of vaccination agreement (OR 1.089 p-value=0.047, OR1.395 p-value=0.003) respectively. On the other hand, respondents who previously developed COVID-19 were significantly more worried about the Monkeypox disease (1.30 times more, p-value=0.020).

**Conclusion:** Worry levels amongst the public are higher from COVID-19 than Monkeypox. Perception of Monkeypox as a dangerous and virulent disease, worry from contracting the disease, and high commitment to infection precautionary measures were predictors of agreement with Monkeypox vaccination. While advanced age and high education level are predictors of low agreement with vaccination.

## 1 Introduction

The emergence of the Monkeypox disease in May 2022 across the globe has caused significant worry among the public, especially with the still ongoing Coronavirus Disease 2019 (COVID-19) pandemic(1). It has previously been demonstrated that the level of worry and concern about the evolving COVID-19 has been exemplified by the different variants, such as the Delta and Omicron(2, 3); therefore, the co-emergence of a new virus potentially might complicate the anxiety and worry levels across the different sectors of the public.

The Monkeypox virus is an enveloped DNA virus, that is a member of the genus Orthopoxvirus, which includes the Smallpox virus (variola major) that was eradicated globally in 1980 by mass vaccination, and this may have resulted in the increased numbers of MOXV cases due to the loss of the partial protective activity of the Smallpox vaccine, due to halt of mandatory vaccination(4).

Recently, Monkeypox disease has increased among travelers to Western and Central African countries, resulting in secondary transmissions from person-to-person(5, 6). Coinciding with the surge of the Omicron variant of COVID-19, the World Health Organization reported the occurrence of 92 laboratory-confirmed Monkeypox cases from the UK, Europe, Australia, Canada, and the USA, mainly among homosexual males. Currently, there are two potential vaccines that could be used in the case of MOXV infection: ACAM2000 orthopoxvirus and JYNNEOS vaccines(7, 8).

With the emergence of the Monkeypox disease outbreak, we rapidly developed and disseminated a survey to assess the public level of worry from it compared to COVID-19 and their readiness for vaccination against it in the general population of Saudi Arabia.

## 2 Method

### 2.1 Data Collection

A structured questionnaire comprising 21 items was used. The survey tool was adopted from our previously published research on COVID-19 with modifications related to the new Monkeypox outbreak (9-13). The questionnaire was translated into Arabic by an expert from the research team and back translated to English to ensure accuracy. The final survey version was piloted among ten members of the general community for clarity and consistency. Modifications were implemented based on the experts’ recommendations. The questionnaire took eight minutes to complete. The research team experts approved the final version of the survey for language accuracy, clarity, and content validity.

Variables surveyed included their sociodemographic characteristics, levels of worries from Monkeypox compared to COVID-19 and sources of the worries, previous COVID-19 infection status, readiness for Monkeypox disease vaccination, and their knowledge about the Monkeypox disease, which was assessed by nine questions assessing their knowledge of modes of transmission and period of contagiousness, the effectiveness of smallpox and chickenpox vaccines against Monkeypox disease.

Also, we assessed their compliance with precautionary measures against COVID-19, and finally, their Generalized Anxiety Disorder (GAD7) score(14, 15). GAD7 is a self-reported, 7-item validated scale. Respondents indicated how frequent they were bothered during the previous two weeks by symptoms of feeling nervous, not being able to stop worrying, worrying about different things, trouble relaxing, restless, irritable, and afraid that something awful may happen. Responses were “not at all,” “several days,” “more than half the days,” and “nearly every day,” and scored as 0, 1, 2, and 3. GAD7 score of ≥10 identified cases of anxiety, with 89% sensitivity and 82% specificity, good internal consistency (Cronbach α□=□.92), and test-retest reliability (intraclass correlation□=□0.83)(14). GAD7 scores were totaled and classified as minimal (0–4), mild (5–9), moderate (10–14), and severe (15–21).

An online, cross-sectional survey of adults in the Kingdom of Saudi Arabia (KSA) was conducted over ten days (from May 27, 2022, until June 5, 2022). Participants were invited by convenience sampling techniques through various social media platforms (Twitter and WhatsApp groups) and email lists. Participants were asked to complete the online survey through the SurveyMonkey© platform, with each response allowed once from each unique IP address to ensure single entries. The first page of the survey explained the research objectives and assured confidentiality.

### 2.2 Ethical Approval

Participants were informed of the purpose of the study, and their voluntary participation was obtained at the beginning of the electronic survey. Ethical approval was granted by the institutional review board (IRB) at King Saud University (22/0416/IRB) before the collection of data.

### 2.3 Statistical analysis

Means and standard deviations were used to describe continuous variables and frequencies and percentages for categorically measured variables. The histogram and the Kolmogorov-Smirnove test were applied to test the assumption of normality, and the Levene’s test was used to test the homogeneity of variance statistical assumption. Cronbach’s alpha test was used to assess the internal consistency of the measured questionnaires. The Multivariate Binary Logistic Regression Analysis was used to assess what factors could explain people’s worry over Monkeypox and their support for vaccination against Monkeypox. The association between predictors with the outcome dependent variables in the multivariate Logistic Binary regression analysis was expressed as a multivariate-adjusted Odds Ratio (OR) with their associated 95% confidence intervals. The SPSS IBM statistical analysis program was used for statistical data analysis(16). The statistical Alpha significance level was considered at 0.050 level.

## 3 Results

The online survey was completed by 1546 individuals. Table-1 displays the descriptive analysis of participants’ sociodemographic characteristics. Most participants were 44 years and under (65%) and had a university degree (64.9%). Almost half of the respondents (46.1%) were employed, with the most reported household monthly income (HHI) between 10,000 and 15,000 Saudi Riyals (2,667.4,000 USD).

**Table-1:**
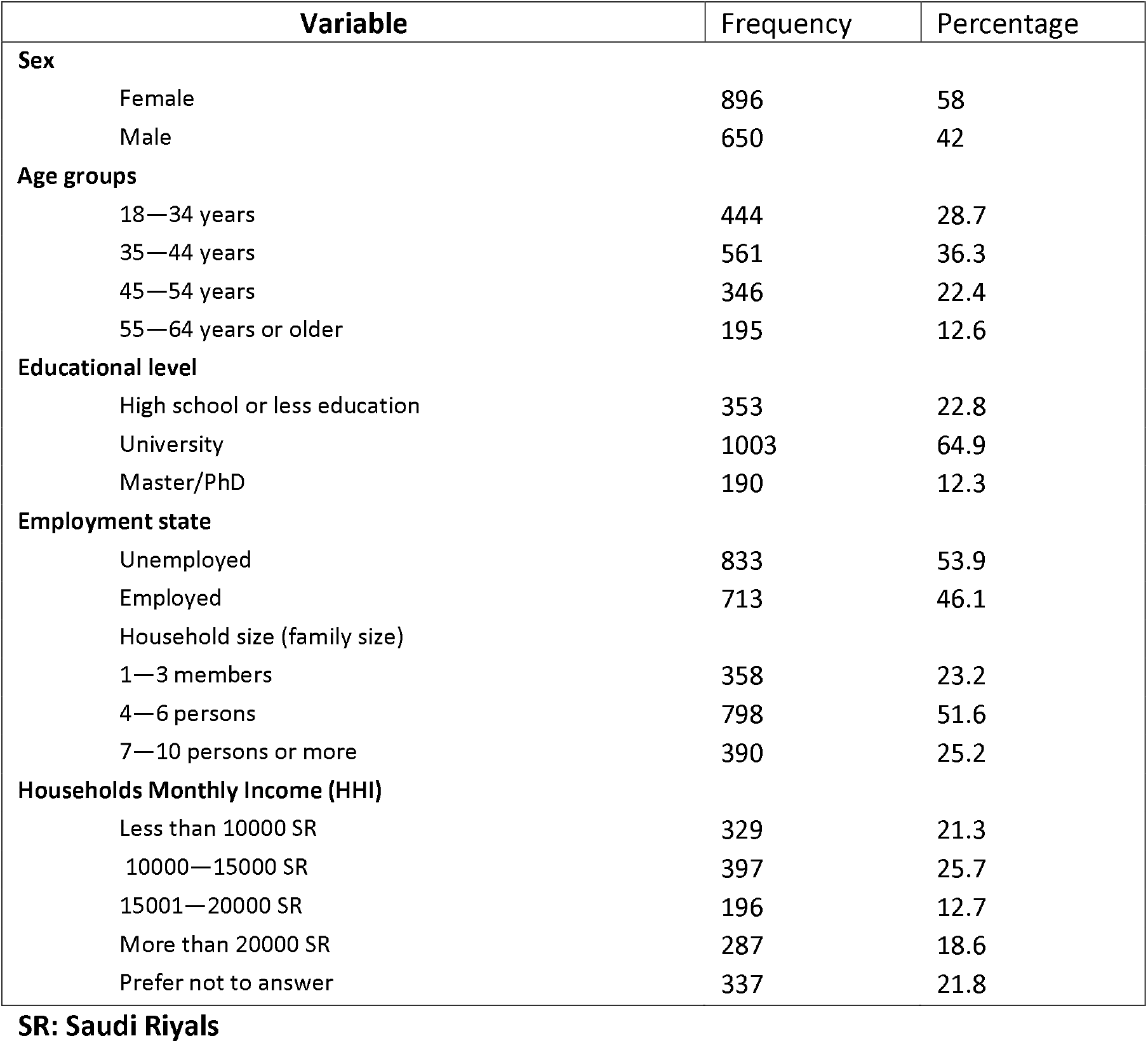
Sociodemographic characteristics of participants (N=1546).

About half of the respondents (49.8%) had previously been infected with COVID-19. Participants’ and family members’ compliance with COVID-19 pandemic precautions revealed that 8.5% were rarely committed, while 48.9% were always committed. The results showed that 62.6% of respondents perceived the COVID-19 pandemic as more worrying than Monkeypox, while 37.4% indicated they were more worried about Monkeypox. More than half of respondents (51.3%) perceived Monkeypox as a dangerous and virulent disease warranting a rapid call for respiratory and contact precautions, as shown in Table 2.

**Table-2:**
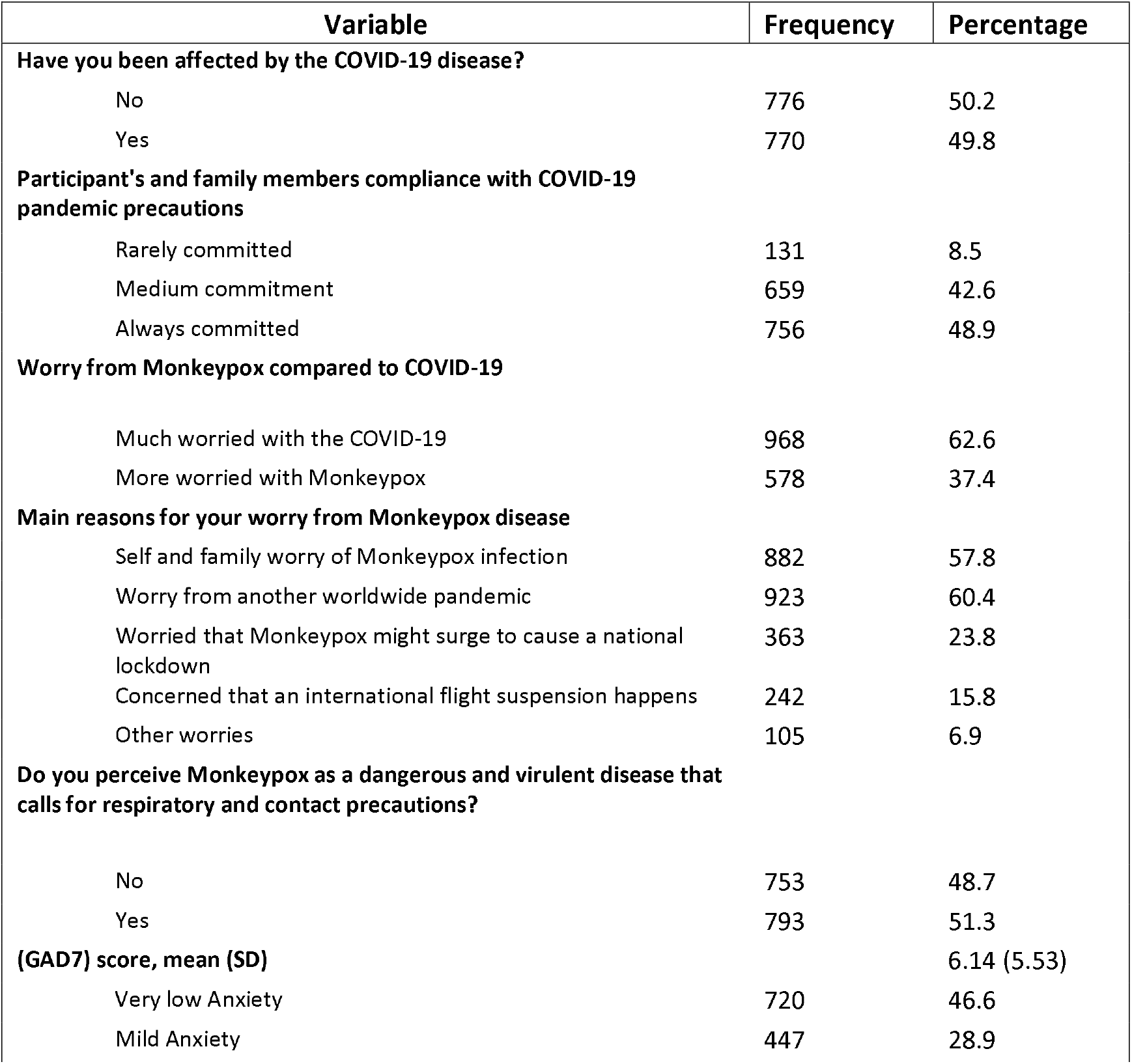
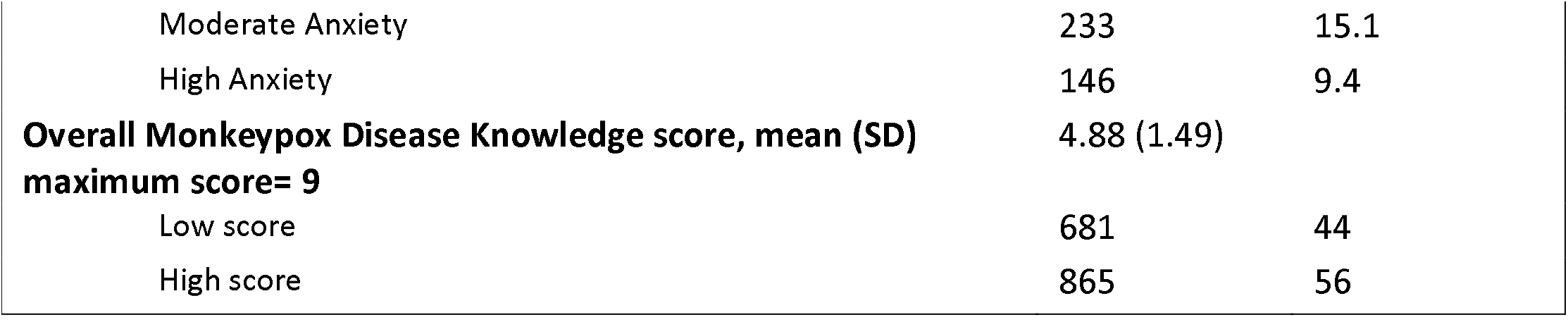
Respondents’ attitudes, perceptions, and beliefs about Monkeypox disease (N=1546).

The average knowledge score from the nine questions assessing respondents’ knowledge regarding Monkeypox was used as a cutoff point to categorize the respondents’ knowledge scores into high or low. The average knowledge score was 4.88 out of 9 SD 1.49, with 56% of respondents achieving a high score. Respondents indicated that they sought information from an official local and international sources, social networks, and other websites.

Respondent’s Overall Generalized Anxiety was measured with the GAD7 scale, their overall mean score was 6.14/21 SD 5.53 points highlighting mild anxiety levels. Table 2

### Worries from Monkeypox Disease and vaccination readiness

Most respondents (60.4%) were worried about the progression of the disease into a global pandemic (figure A). This was followed by worry about themselves and their family’s contracting the disease (57.8%), followed by worry about national lockdown (23.8%) and domestic and international flight shutdowns (15.8%). A minority of respondents (6.9%) perceived other worries from the Monkeypox disease, such as the disfiguring scarring, risks to children and pregnant women, risks of travel restrictions, or requirements of additional mandatory vaccines.

**Figure-A:**
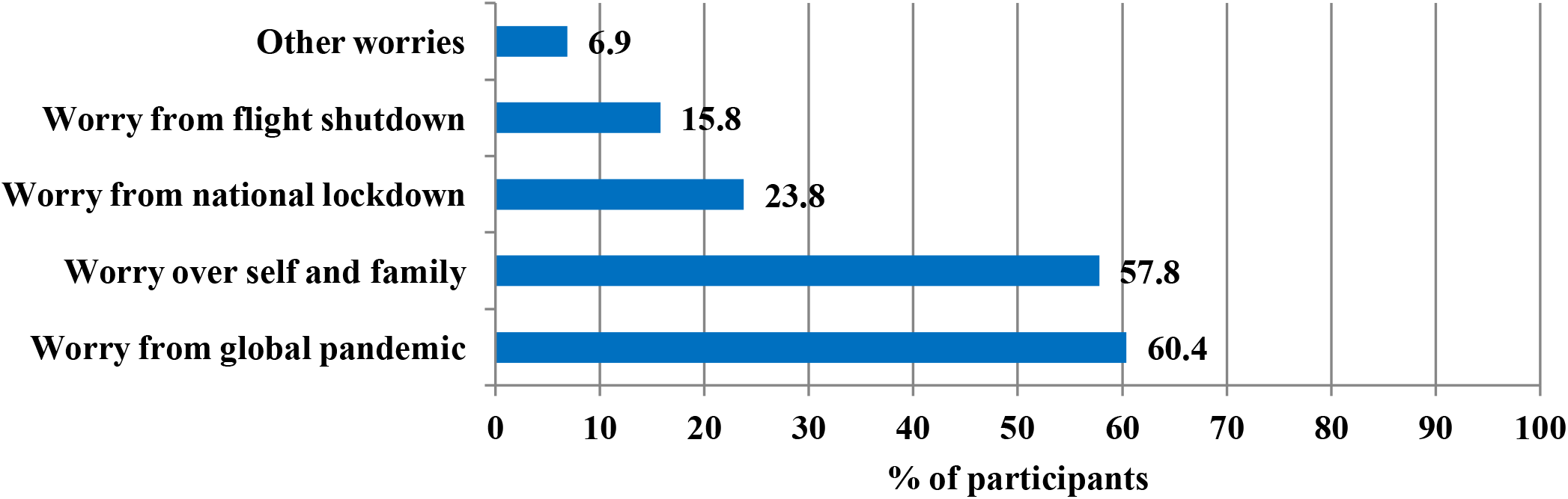
Respondent’s worries from the MonkeyPox Disease.

### Respondents’ agreement with Monkeypox vaccination

In relation to the agreement with Monkeypox vaccination in the current stage of the disease, almost half of respondents (50.6%) favor vaccination. In response to whom participants thought should be vaccinated against Monkeypox disease, 77.7% of respondents felt that immunodeficient persons should be vaccinated, followed by the elderly (62.1%), young children (61.1%), chronically ill patients (59.7%), and healthcare workers (59.4%) (Figure-B).

**Figure-B:**
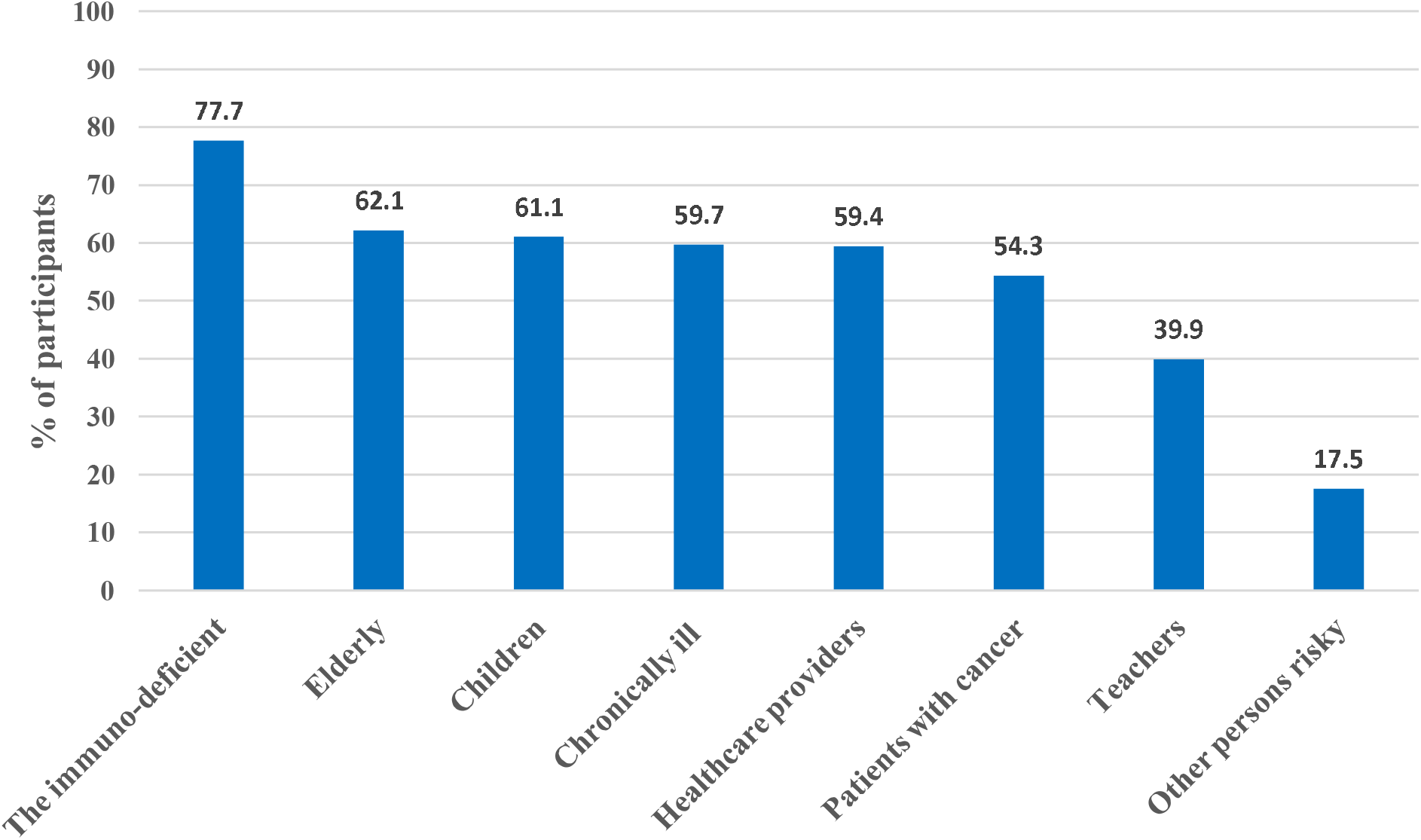
Respondent’s perceptions of MonkeyPox vaccine candidates.

As shown in Table 3, the Multivariate Binary Logistic Regression Analysis identified variables associated with higher odds of respondents’ agreement with vaccination against Monkeypox in the current stage of the disease. Respondents aged 45 years and over were significantly less likely to support vaccination against Monkeypox (OR 0.871, p = 0.006). Respondents who perceived Monkeypox disease as dangerous and virulent were 45.6% more likely to support vaccination p = 0.001

**Table-3:** Multivariate Binary Logistic Regression Analysis of people’s odds of agreement to vaccinate against Monkeypox Disease.

|  | Multivariate adjusted Odds Ratio (OR) | 95% C.I. for (OR) |  | p-value |
| --- | --- | --- | --- | --- |
|  |  | Lower | Upper |  |
| Age group<br>>=45 years | .871 | .790 | .961 | .006 |
| Perceives Monkeypox as dangerous & virulent | 1.456 | 1.165 | 1.820 | .001 |
| Educational Level<br>University degree or higher education | .719 | .618 | .838 | <0.001 |
| Households' monthly income | .959 | .893 | 1.031 | .258 |
| Knowledge score of Monkeypox | 1.048 | .974 | 1.127 | .210 |
| Higher Family compliance level to precautions to COVID-19<br>Moderate to high compliance | 1.089 | 1.001 | 1.184 | .047 |
| Self and family worry from contracting Monkeypox infection | 1.395 | 1.121 | 1.735 | .003 |
| (GAD7) Score | 1.053 | .945 | 1.173 | .349 |
| Read and search more about Monkeypox after the survey | 1.629 | 1.310 | 2.026 | <0.001 |
| More worried from Monkeypox compared to COVID19 | 1.391 | 1.109 | 1.746 | .004 |
| Relies on MOH as a source of Monkeypox information | 1.282 | 1.032 | 1.592 | .025 |
| Relies on other internet sources for Monkeypox information | 1.543 | .904 | 2.633 | .112 |

**DV= agreement to vaccinate against Monkeypox Disease**

Additionally, those who perceived Monkeypox to be more worrisome than COVID-19 had higher odds of agreeing to vaccinate against Monkeypox (OR 1.391 p=0.004). We analyzed the correlation between age and the mean predicted probability of agreement with vaccination against Monkeypox in the two groups of respondents, those who perceived Monkeypox as a dangerous and virulent disease and those who didn’t. In general, increasing age was associated with decreased probability of agreement with vaccination (Figure D). Still, the probability was higher for those who perceived Monkeypox as a dangerous and virulent disease, while those aged 55 years and above and who perceived Monkeypox as a virulent disease had a close probability to those who did not.

**Figure-D:**
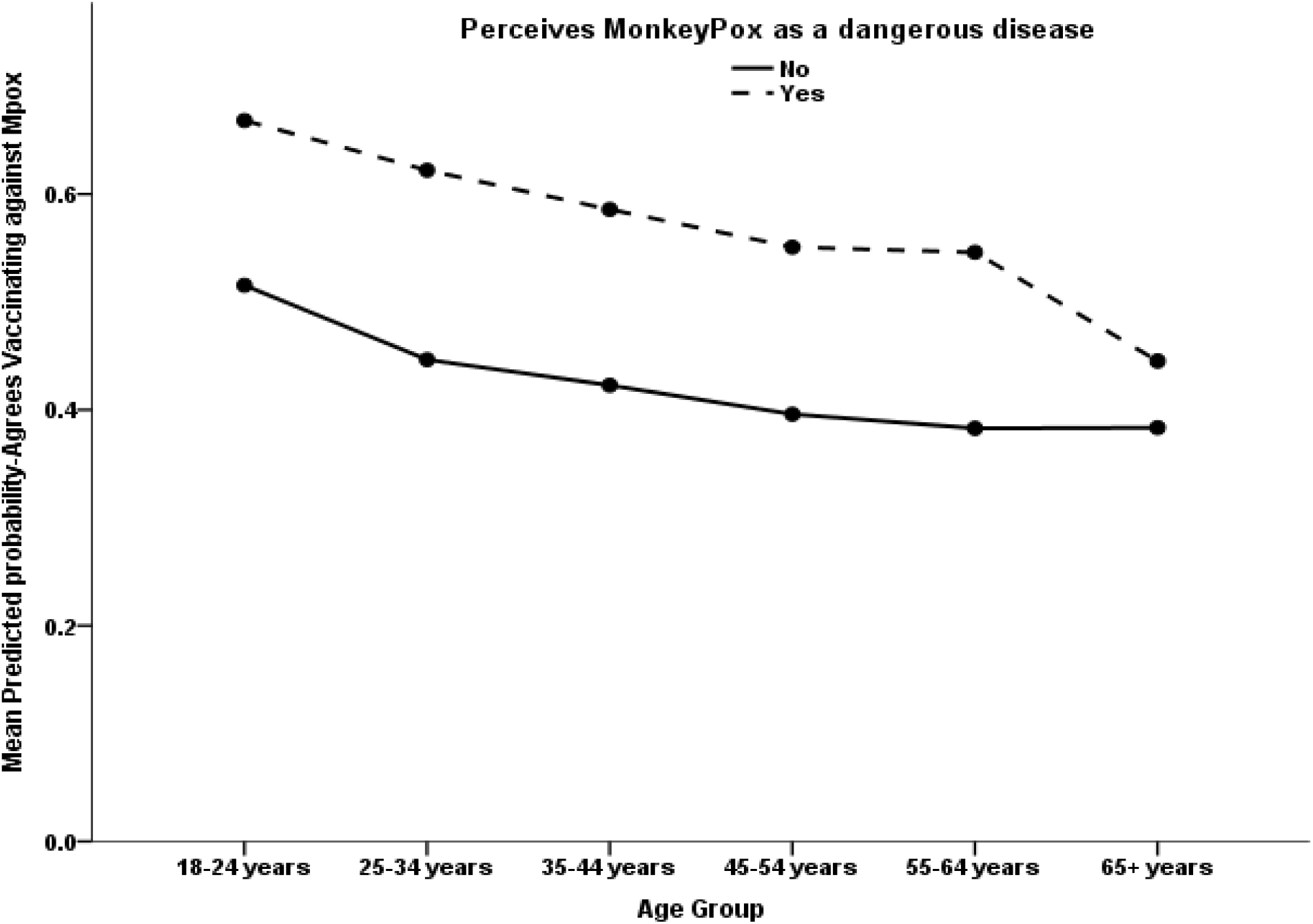
The association between people’s age with their mean predicted probability of supporting the Monkeypox disease vaccines accounting for their perceived danger of Monkeypox.

Those who held university degree or higher had significantly lower odds of supporting vaccination in the current stage (28.1% less OR .719, p <0.001). Respondents’ moderate to high level of self and family commitment to infection control precautionary measures correlated significantly and positively with their agreement to receive Monkeypox vaccination (OR 1.089 p=0.047). Additionally, self and family reported worry about Monkeypox infection were significant predictors of agreement with Monkeypox vaccination (OR 1.395 p=0.003), and respondents who showed greater interest in reading and searching further about Monkeypox had higher odds of supporting vaccination (OR 1.629 p-value<0.001). Those who used the (MOH) website as a source of information compared to those who used other sources of information were significantly more likely to agree with vaccination (OR 1.28 p=0.025). No significant correlation was found between the knowledge score of Monkeypox and the (GAD7) measure of anxiety regarding the agreement to vaccinate against Monkeypox in the current stage.

We assessed variables associated independently with higher worry levels about Monkeypox disease compared to COVID-19 using multivariate logistic regression (Table-4). Age, educational level, employment status, and Monkeypox knowledge score, all did not reveal any significant associations. Participants who perceived Monkeypox as highly dangerous and virulent were found to be significantly more worried about it compared to COVID-19 (OR 3.61, p-value<0.001). It was found that those with moderate to high compliance with infection control precautions were significantly less likely (17.3%) to experience high levels of worry about Monkeypox (OR=0.827 p<0.001). While those who agreed with the Monkeypox vaccination were significantly more worried about it (OR=1.372, p=0.006). Those who were worried about themselves or their families from contracting the infection had slightly higher odds of being more worried; however, this was not statistically significantly (OR 1.245, p=0.066). On the other hand, respondents who previously had COVID-19 were found to be significantly more worried from the Monkeypox disease compared to COVID-19 (OR=1.30 p=0.020).

**Table-4:** Multivariate Binary Logistic Regression Analysis of respondents’ odds of higher worry level from Monkeypox disease compared to COVID-19.

|  | Multivariate adjusted Odds Ratio (OR) | 95% C.I. for (OR) |  | p-value |
| --- | --- | --- | --- | --- |
|  |  | Lower | Upper |  |
| Age group | 1.000 | .903 | 1.109 | .994 |
| Educational Level | .918 | .779 | 1.082 | .308 |
| Households' monthly income | 1.061 | .984 | 1.143 | .124 |
| Employed | .866 | .685 | 1.095 | .231 |
| Monkeypox Knowledge score | .974 | .903 | 1.050 | .488 |
| Perceives Monkeypox as dangerous & virulent | 3.609 | 2.863 | 4.551 | <0.001 |
| Family compliance with universal precautionary level to COVID-19 Moderate or high | .827 | .757 | .903 | <0.001 |
| Agreement with Monkeypox vaccination | 1.372 | 1.095 | 1.719 | .006 |
| Self and family worry from contracting Monkeypox infection | 1.245 | .986 | 1.573 | .066 |
| Previous exposure to COVID-19 Disease Yes | **1.300** | 1.042 | 1.623 | .020 |
| (GAD7) Score | 1.051 | .939 | 1.176 | .385 |

**DV= higher worry level from Monkeypox disease compared to COVID-19.**

## 4 Discussion

During the last two years, the world faced the burden of the COVID-19 pandemic. Though it was not the largest pandemic in the modern history, it had peculiarity due to its occurrence during an era of broad transportation networks and interaction among all nations, concurrently with great development in infection prevention measures compared to previous pandemics, and sophisticated levels of information technology and telecommunication(17). The pandemic caused major disruption for humans, at least health and economic levels. Therefore, with the recent recovery of the international systems from the pandemic consequences and loosening of the tight measures that were applied on travelling and personal communication, the announcement of the reemergence of the Monkeypox disease might potentially have detrimental effect on the societies regarding worry and anxiety(1). The available information about Monkeypox had not yet gained international concern. However, 60.4% of our participants indicated their higher worry about the current Monkeypox virus outbreak becoming a global pandemic. This worry is probably related to their recent experience with the COVID-19 pandemic.

In line with the current COVID-19 pandemic, it is assuring to note that 42.8% of our participants had medium and 48.9% had constant compliance with infection control measures. This is an important aspect for the continued messaging and education that authorities could build on as we go through the alert of Monkeypox. In a commentary, authors called for calls for immediate long-term public and travel health planning(18). The main transmission route of Monkeypox is close and extended contacts with infected individuals(19). Thus, the current precautions in place are of paramount to build on for future plans and interventions.

Vaccination against Monkeypox in the current stage of the disease is a challenging decision for healthcare policy makers, international health societies and local authorities. Public perception regarding the need and acceptance of such decision if taken, for specific groups of the population or most of the population like what happened with COVID-19 deserves studying at the current stage considering the COVID-19 vaccine hesitancy the public went through. Especially after receiving 2 doses of COVID-19 vaccines and the emergence of the Omicron variant with its mild disease nature and questionable vaccines effectiveness against it and other variants. Our cohort has shown that those aging 45 years and above were significantly less supporting vaccination against Monkeypox, the same phenomenon has been reported in public with COVID-19 vaccine even after 2 years of the pandemic.

While public perception of the Monkeypox significantly affected their agreement with vaccination, those who perceived Monkeypox disease as dangerous and virulent (51.3%), Self and family worry of Monkeypox infection (57.8%) and those who perceived it more worrisome compared to COVID-19 (37.4%) had higher odds of agreement with vaccination. That mirrors others work in that regard as perceiving the disease severe was associated with increased willingness for vaccination(20-24). Furthermore, respondents who had moderate to high level of self & family commitment with infection control precautionary measures (91.5%) also showed significant agreement with vaccination against Monkeypox which echoes their healthy infection prevention behavior and is similar to what was observed with COVID-19 vaccines(25). Respondents eager to read about the Monkeypox disease also was a significant predictor of vaccination agreement which translates their perception of disease risk, complacency and collective responsibility toward their families and society.

Meanwhile, only 37.4% of our cohort were more worried of the reemerging Monkeypox compared to COVID-19, 51.3% perceived Monkeypox as a dangerous and virulent disease. Those who perceived it as this had significant worry from it compared to COVID-19. The nature of perception of any disease affects public worry, fear and anxiety levels as all these cognitive attitudes are interconnected(26). On the other hand, those who had moderate to high compliance with infection control precautions (91.5%) had significantly less worry from Monkeypox, which is an expected feeling having decent self-protective behavior that feeds their cognition with safety feelings from remerging infectious diseases(27). That did not echo the healthy behavior of the respondents who agreed with vaccination against Monkeypox (50.6%), who had significant higher worry levels from it. Seeking vaccination may correlate with higher worry levels generally(21, 28). Almost half of our cohort had developed COVID-19 already at the time of the survey, and the majority had a minor disease, even though they had significantly higher worry levels from Monkeypox disease compared to COVID-19, that high worry level can be understood considering the high rates of PTSD, higher perceived life threat and anxiety in COVID-19 survivors(21, 29, 30), while those who had self and family worry from contracting Monkeypox infection had only slight high worry levels from Monkeypox that did not reach statistical significance.

### 4.1 Study limitations and strengths

Our research is subject to the limitation of cross-sectional surveys, such as convenience sampling, variable response rates, and possible recall biases. While this study is among the first to explore perceptions and worries among the public considering the newly reported cases of Monkeypox outside African countries, the respondents’ experiences and perceptions are likely to change as the situation evolves. Moreover, it is noteworthy that there were no cases of Monkeypox reported in Saudi Arabia at the time of data collection, although it was reported in neighboring countries, like the UAE and Jordan. As perceptions may differ from one setting to another, future research could explore this further.

## 5 Conclusions

Worry levels amongst the public were higher regarding COVID-19 than Monkeypox during the first month of the WHO alert about emerging cases of Monkeypox in several countries outside Africa. Perception of Monkeypox as dangerous and virulent disease, worry from contracting the disease and high commitment with infection precautionary measures were predictors of agreement with Monkeypox vaccination. While advanced age and high education level are predictors of low agreement with vaccination.

## Data Availability

All data produced in the present study are available upon reasonable request to the corresponding author.

## Abbreviations

CDC: Centers for Disease Control and Prevention
COVID-19: Coronavirus disease 2019
MOH: Ministry of Health
MOXV: Monkeypox virus
SARS-CoV-2: Severe acute respiratory syndrome coronavirus 2
WHO: World Health Organization

## Acknowledgments

The authors extend their appreciation to the Deanship of Scientific Research, King Saud University, for funding through Vice Deanship of Scientific Research Chairs; Research Chair of Prince Abdullah Ben Khalid Celiac Disease; Riyadh, Kingdom of Saudi Arabia. The research team is thankful for the statistical data analysis consultation offered by www.hodhodata.com.

